# Socioeconomic position and cardiometabolic risk factors in children and adolescents with obesity: findings from the Bern Obesity in Childhood and Adolescence Biorepository (BOCAB)

**DOI:** 10.64898/2026.09.24.26362616

**Authors:** Alexander Koch, Cristina Giachino, Andrew O Agbaje, Markus Juonala, Costan G Magnussen, David P Burgner, Danielle K Longmore, Lorenz M Leuenberger, Claudia E Kuehni, Sibylle Tschumi, Matthias V Kopp, Marco Janner, Christoph Saner

**Affiliations:** Department of Paediatrics, Inselspital, Bern University Hospital, University of Bern, Switzerland; Graduate School for Health Sciences, University of Bern, Switzerland; Institute of Public Health and Clinical Nutrition, School of Medicine, Faculty of Health Sciences, University of Eastern Finland, Kuopio, Finland; Division of Medicine, Turku University Hospital, Turku, Finland; Department of Medicine, University of Turku, Turku, Finland; Alliance for Research in Exercise, Nutrition and Activity (ARENA), School of Allied Health and Human Performance, Adelaide University, Adelaide, South Australia, Australia; Centre for Population Health Research, University of Turku and Turku University Hospital, Turku, Finland; Research Centre of Applied and Preventive Cardiovascular Medicine, University of Turku, Turku, Finland; Murdoch Children’s Research Institute, The Royal Children’s Hospital, Parkville, Victoria, Australia; Department of Paediatrics, University of Melbourne, Parkville, Victoria, Australia; Department of Endocrinology, The Royal Children’s Hospital, Parkville, Victoria, Australia; Institute of Social and Preventive Medicine, University of Bern, Bern, Switzerland; Division of Paediatric Nephrology, Department of Paediatrics, Inselspital, Bern University Hospital, University of Bern, Bern, Switzerland; Division of Paediatric Respiratory Medicine and Allergology, Department of Paediatrics, Inselspital, Bern University Hospital, University of Bern, Bern, Switzerland; Division of Paediatric Endocrinology, Diabetology and Metabolism, Department of Paediatrics, Inselspital, Bern University Hospital, University of Bern, Switzerland

**Keywords:** Childhood obesity, severe obesity, socioeconomic position, Switzerland, cardiometabolic risk, dyslipidemia

## Abstract

**Purpose:** In high-income countries, lower socioeconomic position (SEP) is linked to higher childhood obesity prevalence but its association with cardiometabolic risk in those presenting in pediatric weight management services remain underexplored.

**Methods:** In the Bern Obesity in Childhood and Adolescence Biorepository (BOCAB), we used the Swiss neighborhood index of SEP (0–100, 100=highest SEP) to assess cross-sectional associations with obesity severity and cardiometabolic risk factors, mediation by behavioral lifestyle factors and longitudinal associations between SEP and percentage of the 95^th^ BMI percentile (%BMIp95) and waist circumference-to-height ratio (WHtR), adjusted for age and sex.

**Results:** Among 886 participants (52% male, mean age 11.2 years [SD 3.1]), mean %BMIp95 was 124% (SD 17%). A 10-unit higher SEP was associated with lower %BMIp95 (β –2.57; 95% CI –3.84 to –1.31) and lower WHtR (β –0.006; –0.010 to –0.001), both partially mediated by weekly physical activity and screen time. Higher SEP was also associated with lower odds of insulin resistance (OR 0.80; 0.65 to 0.98), low HDL-cholesterol (OR 0.81; 0.67 to 0.98), elevated non-HDL-cholesterol (OR 0.74; 0.59 to 0.92) and elevated triglycerides (OR 0.83; 0.70 to 0.98). Among 419 participants with longitudinal data, annual reduction in %BMIp95 and WHtR did not differ by SEP.

**Conclusion:** Lower SEP is associated with greater obesity severity and cardiometabolic risk among children and adolescents with obesity presenting at specialized tertiary care. Screening and early referral of lower-SEP children and adolescents with obesity should be encouraged, before cardiometabolic risk accumulates, particularly since treatment response did not differ once enrolled.

## Introduction

Childhood obesity is a global health crisis and is a major risk factor for later cardiovascular and metabolic (cardiometabolic) disease and premature mortality [1, 2]. The prevalence of pediatric obesity in Switzerland is estimated between 3.2% and 6% [3–5], but routine population surveillance fails to differentiate between obesity and severe obesity (defined as BMI >120% of the 95th percentile according to CDC growth charts [6]). This distinction is clinically relevant, as children and adolescents with greater severity of obesity face elevated risks for dyslipidemia, hypertension, insulin resistance, type 2 diabetes mellitus (T2DM) and metabolic dysfunction– associated steatotic liver disease (MASLD) [7].

In high-income countries, lower socioeconomic position (SEP), typically measured by parental education, occupation, or income, is consistently associated with pediatric obesity [8, 9]. Families with lower SEP are often confronted with systemic barriers, including limited access to healthy nutrient-dense foods, contributing to a more obesogenic environment [10]. One study of ∼10,000 children across six European countries found that low SEP was associated with lower vegetable intake, higher consumption of red meat and high-calorie snacks, resulting in nearly four times higher odds of overweight/obesity [11]. The relationship between SEP and physical activity is more nuanced. Evidence suggests that lower-SEP communities have comparable or even greater access to walkable infrastructure and playgrounds, but less access to formal sports facilities [12]— consistent with findings that lower-SEP children engage in less organized physical activity, though evidence on overall physical activity or intensity levels remains mixed [13]. Despite Switzerland’s high ranking in the OECD Better Life Index, there is pervasive health-related socioeconomic inequalities [14]. For example, lower parental education and non-Swiss origin remain consistently associated with obesity risk in the general pediatric population [4, 15–17].

However, evidence on the association between SEP and cardiometabolic health is largely derived from broad, population-based cohorts of predominantly normal-weight children. Large-scale studies, such as ALSPAC from the UK and the multicentric European IDEFICS study, have demonstrated that lower maternal education and SEP are associated with adverse lipid profiles, elevated blood pressure, and higher insulin resistance in childhood [18, 19]. Similarly, the Young Finns Study highlighted that long-term socioeconomic disadvantage correlates with atherosclerosis and fatty liver from childhood into adulthood [20]. While these findings demonstrate a general pattern, there is a scarcity of data regarding the association between SEP and the severity of obesity or the prevalence of cardiometabolic risk factors within high-risk clinical cohorts. One Australian study suggested that family and neighborhood factors are associated with dyslipidemia and liver health in children with severe obesity [21]. Amid growing concern over widening socioeconomic inequalities in high-income countries [22], it remains unknown whether SEP is associated with obesity severity or cardiometabolic risk factors among those who attend a specialized pediatric weight management service, and whether SEP is associated with rate of change in obesity severity under multidisciplinary care.

Addressing this knowledge and translational gap is important to inform risk stratification, promote early referral of high-risk individuals and develop targeted interventions within specialized weight management services.

Therefore, we aimed to investigate the cross-sectional associations between (i) SEP and obesity, including the mediating role of physical activity and screen time, and (ii) SEP and key cardiometabolic risk factors, as well as (iii) the association between SEP and obesity severity over time, in children and adolescents attending a tertiary pediatric weight management service in Switzerland.

## Material and Methods

### Study design and setting

Data were drawn from the Bern Obesity in Childhood and Adolescence Biorepository (BOCAB), an ongoing longitudinal cohort study that has collected data from patients visiting the tertiary weight management service at the Division of Pediatric Endocrinology, University Children’s Hospital Bern since 2011. The baseline assessment consists of three visits covering detailed medical history, diagnostic work-up, and discussion of results, including recommended lifestyle changes and an individualized treatment plan (e.g., dietary counseling, physical therapy, pharmacotherapy). Patients are routinely offered follow-up visits at 6-month intervals until cessation of attendance, obesity resolution, or transition to adult care. A comprehensive cohort profile including a detailed overview of geographic origins of participants and assessments implemented in the study is provided in Online Resource 1 (Figures A.1, A.3 and Table A.1).

Inclusion criteria for this analysis were: (i) age 2–17 years, (ii) attendance at the weight management service at the University Children’s Hospital Bern, and (iii) valid consent defined as either (a) signed BOCAB informed consent or (b) hospital general consent for secondary use of routine health data (any status except declined before 26 April 2022, when the general consent was put in practice; signed or informed but not declined thereafter in accordance with ethics approval by the canton of Bern, BASEC #2022-00968). Exclusion criteria included (i) confirmed or suspected monogenic or syndromic obesity, and (ii) intake of any outcome-modifying medication (e.g., antihypertensive or lipid-lowering agents, metformin, or GLP-1 receptor agonists) at the first clinical attendance (herein termed baseline visit). In addition, participants who initiated GLP-1 receptor agonist therapy or were enrolled in an interventional trial during follow-up were excluded from longitudinal analyses from that time point onward (see Online Resource 1, Figure A.2 for flow diagram of inclusion/exclusion) [23].

### Anthropometry and pubertal status

Height (m) was measured without shoes using a fixed stadiometer to the nearest 0.1 cm. Weight (kg) and body composition—including muscle and fat mass (kg and %)—were assessed in light clothes using a multi-frequency bioelectrical impedance analyzer (InBody770, InBody Co. Ltd., Seoul, Republic of Korea). BMI was calculated as weight divided by height squared (kg/m^2^). Age- and sex-specific reference data from the Centers for Disease Control and Prevention (CDC) were used to calculate BMI standard deviation score (BMI SDS) and percent of the 95^th^ BMI percentile (%BMIp95), with %BMIp95 ≥120% defining severe obesity [20]. %BMIp95 is suggested to provide better discrimination at higher levels of obesity compared to BMI SDS [24]. Waist circumference (WC) was measured at the mid-point between the lateral hip bone and the lower end of the ribs at expiration using a non-flexible tape. The waist circumference-to-height ratio (WHtR) was calculated as WC (cm) divided by height (cm). SDS for WC (WC SDS) were derived from CDC reference data [25]. WHtR provides a reference-curve-independent measure of abdominal adiposity with good performance for cardiometabolic risk screening in children [26–28]. Accordingly, mediation and longitudinal analyses were restricted to %BMIp95 and WHtR as key measures for general and abdominal obesity, respectively. Parental BMI was reported by accompanying parents at first visit. Pubertal status was assessed according to Tanner stages (1–5 with stage 1 defining prepuberty, 2–3 peripuberty, 4–5 postpuberty) [29, 30].

### Cardiometabolic risk factors

Data on cardiometabolic risk factors were assessed by office blood pressure (BP), 24-h ambulatory blood pressure monitoring (ABPM), and fasting venous blood sampling. Office BP was measured by manual auscultation and classified according to the 2023 ESH guidelines [31]; ABPM was performed with validated oscillometric devices and classified according to the 2016 ESH pediatric guidelines with updated 2018 ESC/ESH cut-offs [32, 33]. Blood-based measures comprised the lipid profile (LDL-C, HDL-C, non-HDL-C, triglycerides), glucose metabolism (fasting plasma glucose, HbA1c, and where available 2-h plasma glucose during an OGTT), homeostatic model assessment for insulin resistance (HOMA-IR) and fasting insulin, and alanine aminotransferase (ALT). Continuous biomarkers were analyzed on their original scale and/or as SDS using published pediatric reference data, where indicated. Additionally, binary outcomes (i.e., hypertension, dyslipidemia, prediabetes/suspected T2DM, insulin resistance, elevated ALT) were derived from predefined guideline- or reference-based thresholds for clinical relevance. Full details on devices, quality criteria, reference populations, analytical methods, and all cut-offs are provided in Online Resource 1 (see Extended methods section and Table A.2).

### Family and lifestyle risk factors

Parent-reported family history included prevalent hypertension, dyslipidemia or T2DM among first- or second-degree relatives. Participants reported their average weekly screen time (hours/week) including any exposure to television, laptops, tablets, mobile phones and gaming devices. Participants also self-reported time spent in moderate-to-vigorous physical activity (MVPA, hours/week). MVPA was defined for respondents as activity that noticeably increases breathing and heart rate, consistent with plain-language definitions used in an established self-report instrument [34].

### Swiss neighborhood index of socioeconomic position

We linked the Swiss neighborhood index of socioeconomic position 3 (SSEP 3), an area-based SEP index combining census data from the years 2000 and 2012 to 2015, to the participant’ home address at baseline [35]. This composite measure includes four domains of area-based socioeconomic factors: rent per square meter (as a proxy for income), education level, occupation, and household overcrowding. The Swiss-SEP index was calculated for every residential building in Switzerland by summarizing census data from approximately the nearest 50 households, providing a high-resolution socioeconomic position score for Swiss households [35]. In the following, we refer to this measure as SEP, presented on its continuous scale from 0 (most deprived) to 100 (most advantaged) in descriptive summaries, and rescaled per 10-units for regression and mediation models to facilitate clinical interpretation of effect sizes.

The family’s primary language spoken at home was also recorded and dichotomized into Swiss (German, French, Italian) versus other languages. As direct measures of migration status or race/ethnicity were unavailable, home language was included as a sociodemographic covariate, aiming to partially capture migration-related cultural background and acculturation [36, 37].

### Statistical analysis

Baseline characteristics of study participants are presented as mean (standard deviation [SD]) for continuous variables or number (percentage) for categorical variables. Sex differences were assessed using Welch’s two-sample t-test for continuous variables and chi-square or Fisher’s exact tests for categorical variables. Baseline characteristics were compared between the participants with and without follow-up data to evaluate comparability between subgroups with cross-sectional and longitudinal data (see Online Resource 1, Table A.3).

Cross-sectional associations between SEP (per 10-unit higher SEP) and anthropometric and cardiometabolic outcomes were examined using multivariable linear regression for continuous outcomes and logistic regression for binary outcomes. Regression coefficients and odds ratios with 95% confidence intervals were reported.

Linear models were fitted with robust (heteroscedasticity-consistent) standard errors [38], as some residual plots suggested non-constant variance. Influential observations (Cook’s distance >4/n) were identified; excluding them caused only minor shifts in estimates without consistent direction or pattern across outcomes, so all observations were retained. Covariate adjustments were guided a priori by a directed acyclic graph (DAG, see Online Resource 1, Figure A.4), which identified no covariates required to close backdoor paths between SEP and outcomes [39]. All primary models were adjusted for age and sex. Models additionally adjusted for pubertal stage (Supplementary Model A), home language as candidate proxy for migration background (Supplementary Model B) and home language, parental BMI (maternal and paternal) and, for the relevant cardiometabolic outcomes, the corresponding family history (hypertension, dyslipidemia, or T2DM) (Supplementary Model C) are presented as sensitivity analyses (see Online Resource 1, Table A.4).

Effect modification by sex was assessed for all cross-sectional and longitudinal models by introducing exposure-by-sex interaction terms and comparing model fit via likelihood ratio tests. As no substantial or consistent evidence of sex modification was found across outcomes (data not shown), results are reported for the whole sample throughout.

Mediation analyses were conducted to investigate whether lifestyle factors (i.e., physical activity, screen time) could account for part of the observed associations of SEP with %BMIp95 and WHtR, adjusting for age and sex. Each mediator was assessed individually to decompose the total effect (between exposure [SEP] and outcomes [%BMIp95 and WHtR]) into a direct effect (residual effect without the mediating pathway) and an indirect effect (transmitted through the mediator), based on the counterfactual mediation framework [40] (see Online Resource 1, Figure A.5).

Longitudinal associations between baseline SEP and %BMIp95 and WHtR were analyzed using linear mixed-effects models with participant-specific random intercepts and random slopes for follow-up time to account for repeated measurements within individuals. All available follow-up observations were included, with time since baseline modelled as a continuous variable. Interactions between baseline SEP and follow-up time were tested to determine whether the rate of change in %BMIp95 or WHtR differed by SEP.

All statistical analyses were conducted using R (version 4.4.2; R Foundation for Statistical Computing, Vienna, Austria), including the R package *mediation* [41]. All statistical tests were two-sided. Given the exploratory and hypothesis-driven nature of the analyses, we did not formally adjust for multiple comparisons [42]. Given recommendations against dichotomizing results by statistical significance, we report point estimates with 95% confidence intervals throughout and interpret precision via interval width rather than significance thresholds [43]. An AI language model (Claude Sonnet 5; Anthropic, San Francisco, USA) was used to assist with R coding and guiding statistical analysis, as well as improving the manuscript in terms of readability, consistency and style. All analytic decisions, code, and content were reviewed, verified, and approved by the authors, who take full responsibility for the accuracy and integrity of the work.

## Results

### Characteristics of study population

Data from 886 participants (mean age 11.2 years [SD 3.1], 459 males [51.8%]) were included in the analysis (Table 1). Participants had a mean %BMIp95 of 123.7% (SD 17.4%) corresponding to severe obesity in 471/886 (53.1%). Cardiometabolic risk factor prevalences were as follows: 53/405 (13.1%) had ambulatory hypertension, 383/803 (47.7%) had dyslipidemia, 359/545 (65.9%) had insulin resistance (per HOMA-IR), 171/830 (20.6%) fulfilled biochemical criteria of prediabetes or T2DM, and 381/819 (46.5%) had elevated ALT. Participants’ mean SEP score was 61.1 (SD 9.0), with 358 (40.4%) in the low, 344 (38.8%) in the middle and 184 (20.8%) in the high SEP tertile. Postcode analysis showed participants originated from a wide range of cities beyond the immediate region of Bern (see Online Resource 1, Figure A.1). Swiss languages were the primary language spoken at home for 614/852 participants (72.1%); among these, 566 (92.2%) spoke German, 31 (5.0%) French, and 17 (2.8%) Italian. Sex differences were present across pubertal stages, all anthropometric measures (except unadjusted BMI), 24-h systolic BP (SBP) and 24-h diastolic BP (DBP), FPG, ALT, physical activity and screen time (Table 1). Baseline characteristics of the 419 participants with longitudinal data (median follow-up 18.2 months [IQR 10.3 to 31.5]) were comparable to the 467 participants without follow-up (see Online Resource 1, Table A.3).

**Table 1.** Participant characteristics at baseline, by sex.

| Characteristic | Total<br>(N=886) | Male<br>(N=459) | Female<br>(N=427) | P |
| --- | --- | --- | --- | --- |
| **Demographics** |  |  |  |  |
| Age, years | 11.2 (3.1) | 11.4 (3.1) | 11.0 (3.2) | 0.090 |
| Pubertal stage (Tanner) | [n=776] | [n=391] | [n=385] | **<0.001** |
| Prepubertal | 358 (46.1%) | 217 (55.5%) | 141 (36.6%) |  |
| Peripubertal | 218 (28.1%) | 107 (27.4%) | 111 (28.8%) |  |
| Postpubertal | 200 (25.8%) | 67 (17.1%) | 133 (34.5%) |  |
| SEP | 61.06 (8.95) | 60.92 (8.81) | 61.21 (9.11) | 0.64 |
| Home language | [n=852] | [n=442] | [n=410] | 0.13 |
| Swiss (German/French/Italian) | 614 (72.1%) | 308 (69.7%) | 306 (74.6%) |  |
| Other | 238 (27.9%) | 134 (30.3%) | 104 (25.4%) |  |
| **Anthropometry & body composition** |  |  |  |  |
| Height, cm | 151.5 (17.9) | 153.7 (18.3) | 149.2 (17.1) | **<0.001** |
| Weight, kg | 70.2 (25.6) | 72.6 (26.8) | 67.6 (24.1) | **0.0035** |
| BMI, kg/m<sup>2</sup> | 29.4 (5.5) | 29.5 (5.4) | 29.2 (5.5) | 0.40 |
| BMI SDS | 2.32 (0.61) | 2.36 (0.63) | 2.27 (0.59) | **0.034** |
| %BMIP<sub>95</sub> | 123.7 (17.4) | 125.7 (18.0) | 121.5 (16.5) | **<0.001** |
| WC, cm | 89.1 (13.5) [n=798] | 91.4 (13.8) [n=413] | 86.7 (12.7) [n=385] | **<0.001** |
| WC SDS | 1.78 (0.41) [n=765] | 1.84 (0.40) [n=397] | 1.71 (0.42) [n=368] | **<0.001** |
| WHtR | 0.588 (0.057) [n=798] | 0.595 (0.059) [n=413] | 0.582 (0.055) [n=385] | **0.0013** |
| %BF | 42.0 (6.5) [n=583] | 40.9 (6.6) [n=308] | 43.2 (6.2) [n=275] | **<0.001** |
| %MM | 31.4 (3.6) [n=585] | 32.1 (3.8) [n=309] | 30.5 (3.3) [n=276] | **<0.001** |
| **Blood pressure** |  |  |  |  |
| Office measurements |  |  |  |  |
| Office SBP, mmHg | 109 (12) [n=783] | 109 (12) [n=400] | 109 (11) [n=383] | 0.50 |
| Office DBP, mmHg | 69 (9) [n=781] | 70 (9) [n=399] | 69 (9) [n=382] | 0.39 |
| 24-h ABPM |  |  |  |  |
| 24-h SBP, mmHg | 110 (9) [n=407] | 110 (9) [n=206] | 109 (8) [n=201] | **0.039** |
| 24-h DBP, mmHg | 65 (5) [n=407] | 65 (5) [n=206] | 64 (5) [n=201] | **0.033** |
| **Lipids** |  |  |  |  |
| HDL-C, mmol/L | 1.20 (0.27) [n=812] | 1.20 (0.27) [n=421] | 1.20 (0.27) [n=391] | 0.89 |
| LDL-C, mmol/L | 2.62 (0.72) [n=798] | 2.62 (0.73) [n=410] | 2.62 (0.71) [n=388] | 0.99 |
| Non-HDL-C, mmol/L | 2.90 (0.78) [n=811] | 2.91 (0.79) [n=421] | 2.90 (0.77) [n=390] | 0.76 |
| TG, mmol/L | 1.16 (0.65) [n=811] | 1.16 (0.68) [n=420] | 1.15 (0.61) [n=391] | 0.746 |
| **Metabolism & liver** |  |  |  |  |
| FPG, mmol/L | 5.10 (0.47) [n=826] | 5.16 (0.45) [n=425] | 5.04 (0.49) [n=401] | <**0.001** |
| Fasting insulin, mU/L | 22.4 (16.3) [n=547] | 21.8 (16.2) [n=277] | 23.0 (16.4) [n=270] | 0.39 |
| HOMA-IR | 5.16 (4.25) [n=545] | 5.09 (4.32) [n=275] | 5.24 (4.19) [n=270] | 0.68 |
| 2-h PG, mmol/L | 6.42 (1.38) [n=492] | 6.40 (1.23) [n=263] | 6.45 (1.54) [n=229] | 0.74 |
| HbA1c, % | 5.4 (0.3) [n=348] | 5.4 (0.3) [n=189] | 5.4 (0.4) [n=159] | 0.15 |
| ALT, U/L | 28 (21) [n=819] | 32 (25) [n=422] | 25 (13) [n=397] | <**0.001** |
| **Lifestyle & family** |  |  |  |  |
| Physical activity, h/week | 3.5 (2.6) [n=813] | 3.7 (2.7) [n=413] | 3.3 (2.4) [n=400] | **0.026** |
| Screen time, h/week | 17.7 (12.4) [n=797] | 19.0 (12.5) [n=403] | 16.4 (12.3) [n=394] | **0.0023** |
| Parental BMI |  |  |  |  |
| Maternal BMI, kg/m2 | 28.9 (6.4) [n=671] | 29.1 (6.7) [n=342] | 28.7 (6.1) [n=329] | 0.49 |
| Paternal BMI, kg/m2 | 29.2 (5.0) [n=608] | 29.0 (4.8) [n=308] | 29.4 (5.2) [n=300] | 0.31 |
| Family history of |  |  |  |  |
| T2DM | 385 (49.4%) [n=779] | 197 (48.6%) [n=405] | 188 (50.3%) [n=374] | 0.70 |
| Hypertension | 454 (57.8%) [n=786] | 233 (57.5%) [n=405] | 221 (58.0%) [n=381] | 0.95 |
| Dyslipidemia | 255 (33.4%) [n=763] | 133 (33.9%) [n=392] | 122 (32.9%) [n=371] | 0.82 |

Note. Continuous variables: mean (SD), with N in brackets where less than the column total, compared with Welch's t-test. Categorical variables: n (%) of non-missing, compared with chi-squared or Fisher's exact test.
SEP, socioeconomic position; BMI, body mass index; SDS, standard deviation score; %BMIp95, percentage of the 95th BMI percentile; WC, waist circumference; WHtR, waist-to-height ratio; %BF, percentage body fat; %MM, percentage muscle mass; SBP, systolic blood pressure; DBP, diastolic blood pressure; ABPM, ambulatory blood pressure monitoring; HDL-C, high-density lipoprotein cholesterol; LDL-C, low-density lipoprotein cholesterol; non-HDL-C, non-high-density lipoprotein cholesterol; TG, triglycerides; FPG, fasting plasma glucose; HOMA-IR, homeostatic model assessment for insulin resistance; 2hPG, 2-hour plasma glucose; HbA1c, glycated hemoglobin; ALT, alanine aminotransferase; T2DM, type 2 diabetes mellitus

### Cross-sectional, multivariable regression analyses of SEP with anthropometry, cardiometabolic risk factors and family/lifestyle factors

#### SEP with anthropometry

In age- and sex-adjusted linear regression, higher SEP was associated with lower general obesity, including %BMIp95 (−2.57% per 10-unit higher SEP, 95% CI −3.84 to −1.31, p<0.001); and lower abdominal obesity, including WHtR (−0.006 per 10-unit higher SEP, 95% CI −0.010 to −0.001, p=0.0090) (Table 2). Additional adjustment for pubertal stage or home language (Supplementary Models A and B) did not alter these results, while adjustment for home language and parental BMI (Supplementary Model C) attenuated effect estimates to a small degree including strength of evidence for WHtR, %BF and %MM (see Online Resource 1, Table A.4). Adjusted logistic regression further showed that higher SEP was associated with lower odds of severe obesity (OR 0.78 per 10-unit higher SEP, 95% CI 0.67 to 0.91, p=0.0016) (Table 3).

**Table 2.** Linear regression models of the associations of socioeconomic position (SEP) with anthropometry and cardiometabolic risk factors, adjusted for age and sex. Estimates per 10-units higher SEP.

| Outcome | $\beta$ (95% CI) | p | n |
| --- | --- | --- | --- |
| **Anthropometry & body composition** |  |  |  |
| BMI, kg/m<sup>2</sup> | -0.61 (-0.92, -0.30) | <0.001 | 886 |
| BMI SDS | -0.09 (-0.13, -0.04) | <0.001 | 886 |
| %BMIp95 | -2.57 (-3.84, -1.31) | <0.001 | 886 |
| WC, cm | -1.20 (-1.93, -0.47) | 0.0013 | 798 |
| WC SDS | -0.05 (-0.08, -0.02) | <0.001 | 765 |
| WHtR | -0.006 (-0.010, -0.001) |  | 798 |
| %BF | -0.91 (-1.49, -0.33) | 0.0022 | 583 |
| %MM | 0.50 (0.18, 0.83) | 0.0025 | 585 |
| **Blood pressure** |  |  |  |
| Office measurements |  |  |  |
| Office SBP SDS | -0.05 (-0.13, 0.02) | 0.16 | 783 |
| Office DBP SDS | 0.00 (-0.06, 0.05) | 0.89 | 781 |
| 24-h ABPM |  |  |  |
| 24h SBP SDS | -0.09 (-0.21, 0.03) | 0.16 | 405 |
| 24h DBP SDS | -0.10 (-0.20, -0.01) | 0.034 | 405 |
| **Lipids** |  |  |  |
| HDL-C, mmol/L | 0.01 (-0.01, 0.03) | 0.22 | 812 |
| LDL-C, mmol/L | -0.02 (-0.08, 0.04) | 0.46 | 798 |
| Non-HDL-C, mmol/L | -0.03 (-0.09, 0.03) | 0.31 | 811 |
| TG, mmol/L | -0.04 (-0.09, 0.01) | 0.086 | 811 |
| **Metabolism & liver** |  |  |  |
| FPG, mmol/L | -0.01 (-0.05, 0.02) | 0.46 | 826 |
| Fasting insulin, mU/L | -1.62 (-3.10, -0.15) | 0.031 | 547 |
| HOMA-IR | -0.39 (-0.78, 0.00) | 0.048 | 545 |
| 2-h PG, mmol/L | -0.05 (-0.18, 0.09) | 0.52 | 492 |
| HbA1c, % | -0.03 (-0.08, 0.01) | 0.091 | 348 |
| ALT, U/L | -0.41 (-1.98, 1.16) | 0.61 | 819 |
| **Lifestyle** |  |  |  |
| Physical activity, h/week | 0.24 (0.05, 0.44) | 0.015 | 813 |
| Screen time, h/week | -1.10 (-1.95, -0.25) | **0.011** | 797 |

<sup>a</sup>Sample size reflects complete cases for that model; may vary across models due to missing covariate data.
SEP, socioeconomic position; CI, confidence interval; BMI, body mass index; SDS, standard deviation score; %BMIP95, percentage of the 95th BMI percentile; WC, waist circumference; WHtR, waist-to-height ratio; %BF, percentage body fat; %MM, percentage muscle mass; SBP, systolic blood pressure; DBP, diastolic blood pressure; ABPM, ambulatory blood pressure monitoring; HDL-C, high-density lipoprotein cholesterol; LDL-C, low-density lipoprotein cholesterol; non-HDL-C, non-high-density lipoprotein cholesterol; TG, triglycerides; FPG, fasting plasma glucose; HOMA-IR, homeostatic model assessment for insulin resistance; 2hPG, 2-hour plasma glucose; HbA1c, glycated hemoglobin; ALT, alanine aminotransferase; T2DM, type 2 diabetes mellitus

**Table 3.** Logistic regression models of the associations of socioeconomic position (SEP) with cardiometabolic risk factors, adjusted for age and sex. Estimates per 10-units higher SEP.

| Outcome | OR (95% CI) | p | n<sup>a</sup> |
| --- | --- | --- | --- |
| **Anthropometry** |  |  |  |
| Severe obesity | 0.78 (0.67, 0.91) | **0.0016** | 886 |
| **Blood pressure** |  |  |  |
| Office hypertension | 0.94 (0.76, 1.15) | 0.52 | 778 |
| Ambulatory hypertension (24-h) | 0.82 (0.61, 1.12) | 0.21 | 405 |
| **Lipids** |  |  |  |
| Dyslipidemia | 0.87 (0.74, 1.02) | 0.082 | 803 |
| Low HDL-C | 0.80 (0.66, 0.96) | **0.018** | 812 |
| Elevated LDL-C | 0.89 (0.71, 1.11) | 0.30 | 798 |
| Elevated non-HDL-C | 0.74 (0.60, 0.93) | **0.0080** | 811 |
| Elevated TG | 0.83 (0.70, 0.98) | **0.033** | 811 |
| **Metabolism & liver** |  |  |  |
| Prediabetes/T2DM | 0.89 (0.73, 1.08) | 0.24 | 830 |
| Elevated HOMA-IR | 0.80 (0.65, 0.98) | **0.031** | 545 |
| Elevated ALT | 0.96 (0.83, 1.13) | 0.65 | 819 |

<sup>a</sup>Sample size reflects complete cases for that model; may vary across models due to missing covariate data.
OR, odds ratio; CI, confidence interval; HDL-C, high-density lipoprotein cholesterol; LDL-C, low-density lipoprotein cholesterol; non-HDL-C, non-high-density lipoprotein cholesterol; TG, triglycerides; T2DM, type 2 diabetes mellitus; HOMA-IR, Homeostasis model assessment for insulin resistance; ALT, alanine aminotransferase.

#### SEP with cardiometabolic risk factors

In age- and sex-adjusted linear regression, higher SEP was associated with lower 24-h DBP SDS (−0.10 SDS per 10-unit higher SEP, 95% CI −0.20 to –0.01, p=0.034), while association with 24-h SBP SDS had a comparable point estimate but wider interval (−0.09 SDS per 10-unit higher SEP, 95% CI −0.21 to 0.02, p=0.120). Also, higher SEP was associated with lower fasting insulin (−1.62 mU/L per 10-unit higher SEP, 95% CI −3.10 to –0.15, p=0.031), and lower HOMA-IR (−0.39 per 10-unit higher SEP, 95% CI −0.78 to 0.00, p=0.048) (Table 2). Evidence for an association with triglycerides was similarly directed but less precise (−0.04 mmol/L per 10-unit higher SEP, 95% CI −0.09 to 0.01, p=0.086). Associations with fasting insulin and HOMA-IR were attenuated in all supplementary models (see Online Resource 1, Table A.4). In adjusted logistic regression, higher SEP was associated with lower odds of low HDL-C (OR 0.80 per 10-unit higher SEP, 95% CI 0.66 to 0.96, p=0.018), elevated non-HDL-C (OR 0.74 per 10-unit higher SEP, 95% CI 0.60 to 0.93, p=0.0080), elevated triglycerides (OR 0.83 per 10-unit higher SEP, 95% CI 0.70 to 0.98, p=0.033) and elevated HOMA-IR (OR 0.80 per 10-unit higher SEP, 95% CI 0.65 to 0.98, p=0.031), while the OR for any type of dyslipidemia was comparable in magnitude but less certain (OR 0.87 per 10-unit higher SEP, 95% CI 0.74 to 1.02, p=0.0080) (Table 3).

#### SEP with lifestyle factors

Higher SEP was associated with higher physical activity (0.24 h/week per 10-unit higher SEP, 95% CI 0.05 to 0.44, p=0.015) and lower screen time (–1.10 h/week per 10-unit higher SEP, 95% CI –1.95 to –0.25, p=0.011) (Table 2).

#### Mediation analysis

In age- and sex-adjusted mediation analyses (Table 4), higher weekly physical activity partially mediated associations between SEP and %BMIp95 (indirect effect: –0.21% in %BMIp95 per 10-unit higher SEP via physical activity, 95% CI −0.45 to –0.03, p=0.012; proportion of the total effect mediated: 7.7%) and SEP and WHtR (indirect effect: –0.001 in WHtR per 10-unit higher SEP via physical activity, 95% CI −0.002 to 0.000, p=0.010; proportion of the total effect mediated: 14.9%). Lower screen time partially mediated the association between SEP and %BMIp95 (indirect effect: –0.35% in %BMIp95 per 10-unit higher SEP via screen time, 95% CI −0.68 to –0.05, p=0.020; proportion of the total effect mediated: 12.6%), and SEP and WHtR (indirect effect: –0.001 in WHtR per 10-unit higher SEP via physical activity, 95% CI −0.002 to 0.000, p=0.029; proportion of the total effect mediated: 15.1%).

**Table 4.** Exploratory mediation analysis of the association between socioeconomic position (SEP) and anthropometric outcomes (%BMIp95, WHtR) by behavioral lifestyle factors (physical activity, screentime, fruit/vegetable intake, sweet/salty snack intake), adjusted for age and sex.

| Outcome | Mediator | Indirect effect (95% CI) | p | Direct effect (95% CI) | p | Total effect (95% CI) | p | % mediated<sup>a</sup> | N |
| --- | --- | --- | --- | --- | --- | --- | --- | --- | --- |
| %BMIp95 | Physical activity, h/week | -0.21 (-0.45, -0.03) | **0.012** | -2.51 (-3.79, -1.28) | **<0.001** | -2.72 (-4.02, -1.48) | **<0.001** | 7.7% | 813 |
|  | Screen time, h/week | -0.35 (-0.68, -0.05) | **0.020** | -2.40 (-3.68, -1.15) | **<0.001** | -2.75 (-4.04, -1.47) | **<0.001** | 12.6% | 797 |
| WHtR | Physical activity, h/week | -0.001 (-0.002, 0.000) | **0.010** | -0.005 (-0.009, -0.001) | **0.025** | -0.006 (-0.010, -0.001) | **0.0096** | 14.9% | 752 |
|  | Screen time, h/week | -0.001 (-0.002, 0.000) | **0.029** | -0.005 (-0.009, 0.000) | **0.041** | -0.005 (-0.010, 0.001) | **0.017** | 15.1% | 740 |

<sup>a</sup>ratio of indirect/total effect (i.e., proportion of the total effect explained by the mediator)

#### Longitudinal multivariable mixed-effects regression analysis of SEP with anthropometry

In age- and sex-adjusted longitudinal mixed-effects models (Table 5), higher SEP was associated with lower %BMIp95 and WHtR at baseline (in accordance with linear regression models). Annual change during follow-up was −1.62% per year for %BMIp95 (95% CI −2.30 to −0.93, p<0.001) and −0.006 units per year for WHtR (95% CI −0.009 to −0.003, p<0.001). SEP did not affect the rate of change in %BMIp95 or WHtR (Table 5, and Online Resource 1, Figure A.6).

**Table 5.** Longitudinal mixed-effects models of anthropometric outcomes by socioeconomic position (SEP), adjusted for age and sex. Estimates per 10-unit higher SEP.

| Outcome | Effect | $\beta$ (95% CI) | p | N<sup>a</sup> |
| --- | --- | --- | --- | --- |
| %BMIp95 | Baseline SEP effect | -3.01 (-4.91, -1.12) | **0.0019** | 419 |
|  | Annual change | -1.62 (-2.30, -0.93) | **<0.001** | 419 |
| | SEP × annual change | -0.29 (-1.04, 0.47) | 0.45 | 419 |
| WHtR | Baseline SEP effect | -0.010 (-0.016, -0.004) | **0.0016** | 415 |
|  | Annual change | -0.006 (-0.009, -0.003) | **<0.001** | 415 |
| | SEP × annual change | -0.002 (-0.005, 0.001) | 0.25 | 415 |

<sup>a</sup>N = number of participants with $\geq 2$ visits contributing to that model
Baseline SEP effect = association between SEP and the outcome at baseline; Annual change = mean yearly change in outcome over follow-up period; SEP × annual change = effect modification of the annual trajectory by SEP.
CI, confidence interval; %BMIp95, percentage of the 95th BMI percentile; WHtR, waist-to-height ratio.

## Discussion

In this study, we investigated associations of SEP with obesity severity—including its mediation through behavioral lifestyle factors—and with cardiometabolic risk factors in a large cohort of children and adolescents with, on average, severe obesity attending a Swiss tertiary pediatric weight management service. Our main findings were that (i) higher SEP was associated with lower general (%BMIp95) and abdominal obesity (WHtR), both partially mediated by physical activity and screen time; (ii) higher SEP was associated with a more favorable cardiometabolic risk profile, including lower diastolic blood pressure and markers of insulin resistance and dyslipidemia; and (iii) although higher SEP was associated with less severe obesity at baseline, longitudinal analyses showed that annual reductions in %BMIp95 and WHtR did not differ by SEP.

Our findings align with published associations between higher SEP and lower BMI [44], lower fat mass [45], and cardiometabolic risk factors (dyslipidemia, lower blood pressure) [46, 47]. However, these studies were conducted in general pediatric populations from high-income countries. Our results extend the evidence to children and adolescents with obesity, a particularly vulnerable population for adverse cardiometabolic risk profiles. Comparable results are available from studies in the Australian COBRA cohort, including 444 children and adolescents with severe obesity, where area-based or family-reported SEP variables were associated with BMI, WC, %BF, and liver steatosis [21]. In this study, specific neighborhood characteristics including recreational areas and shopping facilities were associated with dyslipidemia. However, the prevalence of dyslipidemia in the COBRA cohort was much lower (24.4% vs. 47.7% in this study), which limits comparison alongside differences in applied SEP indices, and socio-economic context.

Suggested mechanisms underlying the inverse association between SEP and obesity severity as well as cardiometabolic risk in high-income countries are plausibly related to environmental, nutritional, behavioral and family-related factors. These include the widespread availability of low-cost, energy-dense foods low in nutritional value, high consumption of sugar-sweetened beverages, lower physical activity levels, and higher parental BMI with adoption of unhealthy parental behaviors [8, 11, 48, 49]. In line with these mechanisms, our mediation analysis found that physical activity and screen time each accounted for a modest proportion of the SEP-%BMIp95 and SEP-WHtR associations, suggesting these behaviors partly explain socioeconomic disparities in obesity severity in our cohort. Given the cross-sectional, self-reported nature of the behavioral measures—and the possibility that physical activity or screen time are themselves influenced by a child’s obesity status—these findings are hypothesis-generating rather than evidence of a specific causal mechanism. The remaining, unexplained portion of the SEP-obesity association points to additional pathways, including the food and built environment and unmeasured family or psychosocial factors.

Furthermore, our finding of an inverse association between SEP and diastolic blood pressure is consistent with previous results from both adult and pediatric cohorts [46, 50, 51]. In a prospective Dutch cohort, lower maternal education was associated with higher office blood pressure and increased risk of prehypertension as early as ages 5–6 years, with children’s BMI partly mediating these associations [51]. Findings from the Young Finns Study further showed that early life socioeconomic effects on blood pressure track through adolescence into adulthood [46]. While these studies used office measurements, there is one other study, which examined socioeconomic factors in relation to 24-h ABPM in 212 US adolescents and found associations of neighborhood income with systolic blood pressure, and individual race with diastolic blood pressure [52].Our findings expand on this evidence in a cohort of children and adolescents with severe obesity. Socioeconomic adversity may contribute to elevated blood pressure through pathways involving chronic stress, sympathetic activation, endothelial dysfunction, and vascular changes, potentially amplifying the adverse effects of other stressors such as obesity [53].

Dyslipidemia in children is frequently considered secondary to excessive weight, primarily driven by insulin resistance and alterations in lipoprotein metabolism, such as impaired TG-rich lipoprotein clearance [54, 55].

Comprehensive metabolomics from the COBRA cohort showed that higher %BF was associated with lipid-enriched HDL-C particles (i.e., impaired anti-atherogenic function) and a reduced ratio of docosahexaenoic acid (DHA, atheroprotective via lowering TG and non-HDL-C) to total fatty acids [56]. Similar metabolomic profiles were associated with low SEP (paternal occupation proxy) independent of BMI and diet in 4000 predominantly normal-weight children from the ALSPAC cohort: lower DHA and omega-3 fatty acids, higher monounsaturated fatty acids, and smaller, dysfunctional cholesterol-enriched HDL particles [57]. While these findings raise the possibility of a combined or overlapping effect of obesity severity and low SEP on lipid metabolism, our study cannot distinguish the underlying biological or behavioral pathways involved. Notably, while higher SEP was associated with lower odds of clinically defined low HDL-C, elevated non-HDL-C and elevated triglycerides, continuous linear models for these same lipid measures showed absent or weaker associations (Table 2). This pattern may indicate that SEP-related differences in lipid profiles are concentrated near clinically relevant thresholds rather than reflecting a uniform linear trend across the full distribution, though a modest influence of a small number of influential observations on the continuous estimates, as suggested by our sensitivity analyses, cannot be excluded.

While the direction of association between SEP and insulin resistance in children depends on country-level economic development [58], our findings of higher insulin, HOMA-IR and insulin resistance among participants from lower SEP is consistent with evidence from general pediatric populations from other high-income countries [19, 59]. One longitudinal US study showed that lower parental education is associated with greater insulin resistance and worsening over time, particularly in adolescents with obesity [60]. Prospective data from the IDEFICS study showed that lower parental education was associated with insulin resistance in univariable models, but not after adjusting for BMI SDS or WC SDS, emphasizing the major role of obesity in driving insulin resistance [61]. The central role of adiposity is further evident in the rising prevalence of insulin resistance across obesity severity classes in the NHANES cohort [7]. Although obesity is arguably the main driver of insulin resistance, children and adolescents with obesity from lower SEP likely carry a compound risk.

Overall, the demographic and obesity severity characteristics of the BOCAB cohort are comparable to other cohorts such as the Australian COBRA cohort (mean age 11.1 years, mean BMI SDS 2.5) [21], an Italian cohort (n=453; mean age 11.2 years, mean BMI SDS 2.7) [62], and the German multicenter Adiposity Patients Registry (APV) (n = 65,453; median age 12.8 years, median BMI SDS 2.06) [63]. Prevalences of cardiometabolic risk factors among BOCAB participants (Figure 1) also fall within the range reported in multiple European pediatric obesity studies, where median prevalences were 20.1% for hypertension, 23% for NAFLD, and 48.8% for dyslipidemia [64]. These similarities suggest that BOCAB reflects the demographic and clinical profile typical of children and adolescents in European and Australian specialized/tertiary weight management services, supporting the relevance of our findings to similar clinical populations.

**Fig. 1.**
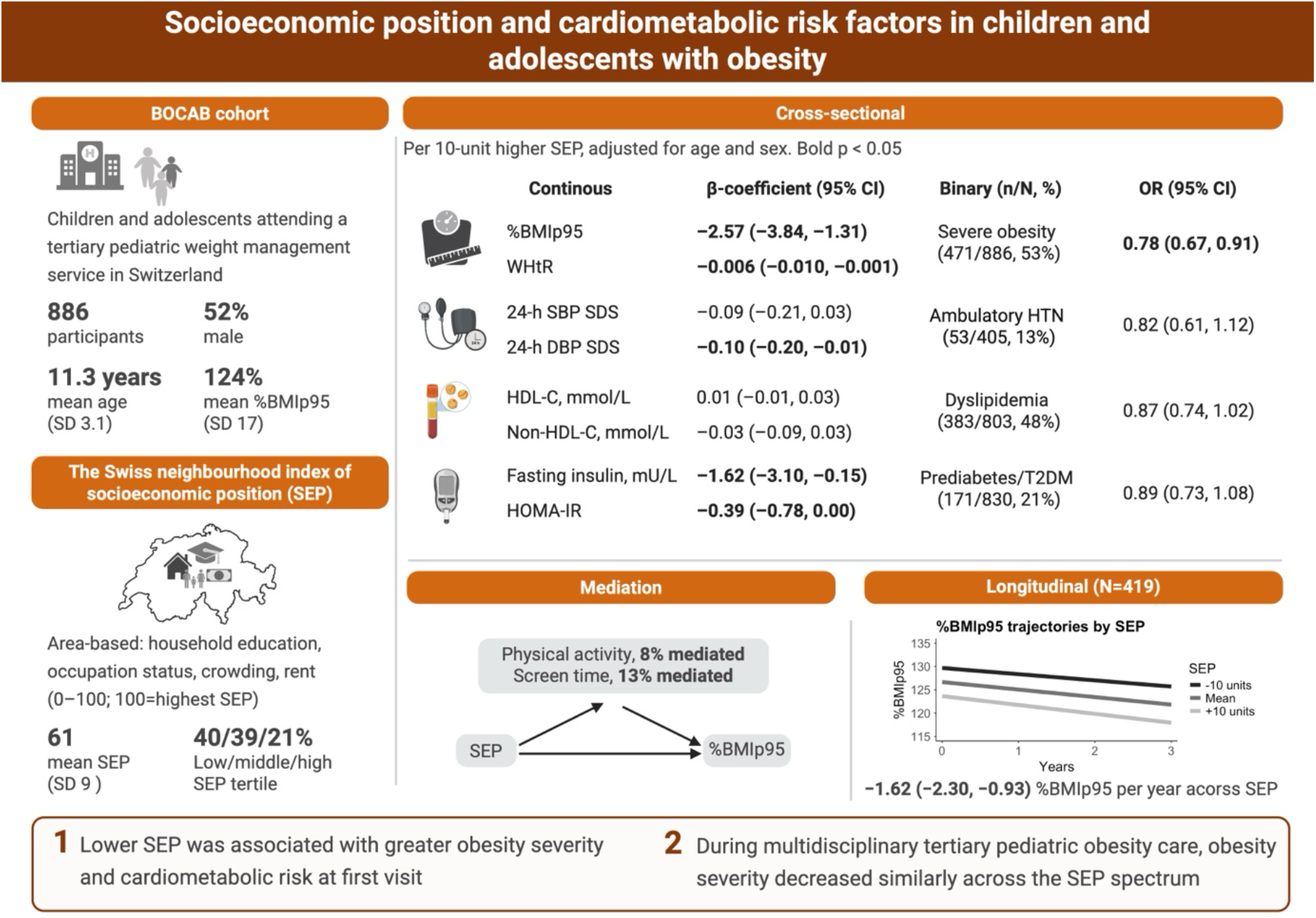
Graphical abstract. All models were adjusted for age and sex. %BMIp95, percentage of the 95th BMI percentile for age and sex; WHtR, waist-to-height ratio; SBP, systolic blood pressure; DBP, diastolic blood pressure; HTN, hypertension; HDL-C, high-density lipoprotein cholesterol; HOMA-IR, homeostatic model assessment for insulin resistance; T2DM, type 2 diabetes mellitus. Created in BioRender. Koch, A. (2026) https://BioRender.com/flzvf0v

Participants in this study achieved a mean annual change in of 1.62% in %BMIp95 and 0.006 units in WHtR, with no evidence that this rate of change differed by SEP. This observation of socioeconomic equity in obesity severity reduction contrasts with findings from the APV cohort, where children from lower SEP backgrounds achieved smaller reductions in BMI compared with their higher SEP counterparts [65]. This discrepancy may reflect differences in program intensity, care model, or population characteristics between the two settings.

Evidence on associations between SEP and changes in obesity measures among children and adolescents with severe obesity remains limited and heterogeneous overall [66], making it difficult to determine whether our findings reflect a feature specific to the Swiss healthcare context, to BOCAB’s multidisciplinary care model, or a broader pattern not yet well captured across similar interventions.

Our study has several strengths, including a large sample of 886 children and adolescents with severe obesity, comprehensive assessment of anthropometric and cardiometabolic parameters and longitudinal tracking of a large subgroup of 419 participants. This study further benefits from an area-based SEP composite aggregating census-level data on rent (proxy for income), education, occupation and overcrowding from participants’ home addresses, reducing missing data and reporting bias compared to self-reports. While our SEP measure shows strong construct validity against self-reported household income data [35, 67], area-based SEP measures also come with some limitations as they risk misclassifying individuals who differ from neighborhood averages.

Accordingly, agreement with self-reports may vary by parameter selection, granularity, and population setting [68]. Additional limitations should be noted. First, the observational, cross-sectional design of the majority of our analyses precludes causal inference regarding the associations between SEP, lifestyle factors, and cardiometabolic outcomes. Physical activity and screen time were assessed by parent- or self-report at a single time point, further limiting causal interpretation of the mediation analyses; longitudinal or objectively measured behavioral data would be needed to clarify the direction and stability of these pathways. We did not perform mediation analysis in our longitudinal subset, since this would be diluted by the effect of the weight management intervention. Second, this study was conducted at a single tertiary referral center; while this enhances internal consistency of assessments and care, it may limit generalizability of our findings to population-based or non-specialized healthcare settings, as well as to healthcare systems structured differently from Switzerland’s. Third, potential attrition bias due to loss to follow-up cannot be excluded, although differences in baseline characteristics of participants with vs. without follow-up were generally small in magnitude (see Online Resource 1, Table A.3), suggesting limited potential for substantial attrition bias. Finally, both residual confounding—for example, from imperfect measurement of home language as a proxy for migration background (Supplementary Model B)—and unmeasured confounding from variables not captured in this study, such as family health beliefs, diet and broader psychosocial factors (e.g., family stress, child or parental mental health), may persist in our analyses.

Our findings have implications for specialized pediatric obesity care. Registry data such as BOCAB capture trends in a high-risk clinical population that is not well represented in general population studies, and our results indicate that socioeconomic disparities in obesity severity and cardiometabolic risk persist even among children already referred for specialized treatment. This underscores the need for clinical programs to actively account for family socioeconomic resources, for example, through subsidized access to healthy foods, sport clubs, and recreational activities for enrolled families, rather than assuming a uniform capacity to implement lifestyle recommendations across all socioeconomic backgrounds. Personalized multidisciplinary lifestyle interventions accounting for family resources can prevent stigmatization, improve outcomes, and even promote upward socioeconomic mobility (e.g., completion of ≥12 years of school) [69]. These children face a dual burden of cardiometabolic risks plus stigmatization, bullying, social isolation and low self-esteem [70]—contributing to a 2.6-fold higher all-cause mortality rate in early adulthood among youth with obesity from low- vs. high-SEP backgrounds [71]. Whether earlier, population-level prevention efforts could reduce the number of children arriving at specialized care with both severe obesity and socioeconomic disadvantage remains an important question for future research.

## Conclusions

This study demonstrates that lower SEP is associated with greater obesity severity and an adverse cardiometabolic risk profile among children and adolescents with obesity presenting at a Swiss tertiary weight management service, with physical activity and screen time only partly explaining the SEP-obesity association. Notably, reduction in obesity severity during treatment remained unaffected by SEP, suggesting that structured multidisciplinary care can be delivered equitably across socioeconomic strata once children are enrolled. These findings highlight the importance of earlier identification and referral of children and adolescents with obesity from lower-SEP families, before cardiometabolic risk factors accumulate, and point to a need for interventions that address underlying socioeconomic barriers to healthy behaviors, alongside intensive individual behavioral and lifestyle counselling.

## Data Availability

All data produced in the present study are available upon reasonable request to the authors.

## Acknowledgements

We are grateful to the BOCAB participants and their families. Special thanks go to the staff of the pediatric weight management service at the University Children’s Hospital Bern and to the PedNet Team Bern for their support with data collection and maintenance.

## Author Contributions

Alexander Koch, Marco Janner, and Christoph Saner contributed to the study conception and design. Material preparation, data collection, and analysis were performed by Alexander Koch and Christoph Saner. The first draft of the manuscript was written by Alexander Koch. All authors commented on previous versions of the manuscript and contributed according to their respective fields of specialty. All authors read and approved the final manuscript.

## Ethics approval

The study was registered in the Registry of All Projects in Switzerland (RAPS; BASEC ID 2022-00968) and approved by the Ethics Committee of the Canton of Bern on 19 August 2022 and follows the ethical guidelines of the 1975 declaration of Helsinki.

## Consent to participate

Written informed consent was obtained from a parent or legal guardian for participants under 18 years of age. Where specific consent was not obtained, inclusion of routine health data was permitted under the general consent for secondary use of routine health data, as approved by the ethics committee.

## Consent to publish

Not applicable.

