## Supplementary Material for "Socioeconomic position and cardiometabolic risk factors in children and adolescents with obesity: findings from the Bern Obesity in Childhood and Adolescence Biorepository (BOCAB)"

Article title:

Department of Paediatrics

### BOCAB – background and objective

The Bern Obesity in Childhood and Adolescence Biorepository (BOCAB) is an ongoing registry with associated biobank designed to collect baseline and longitudinal clinical data and biospecimens from children and adolescents with obesity, as well as non-obese controls. The BOCAB protocol was approved on 18 August 2022 (BASEC-ID 2022-00968) for retrospective inclusion of patients attending the pediatric weight management service at Bern University Children's Hospital since 2011, with prospective enrollment of participants and longitudinal follow-up thereafter (see Figure A.1 for origin of participants and Figure A.2 for inclusion/exclusion flow chart).

The primary objective of BOCAB is to investigate determinants of childhood obesity and the development of related adverse health outcomes thorough comprehensive phenotyping (see Figure A.3 and Table A.1).

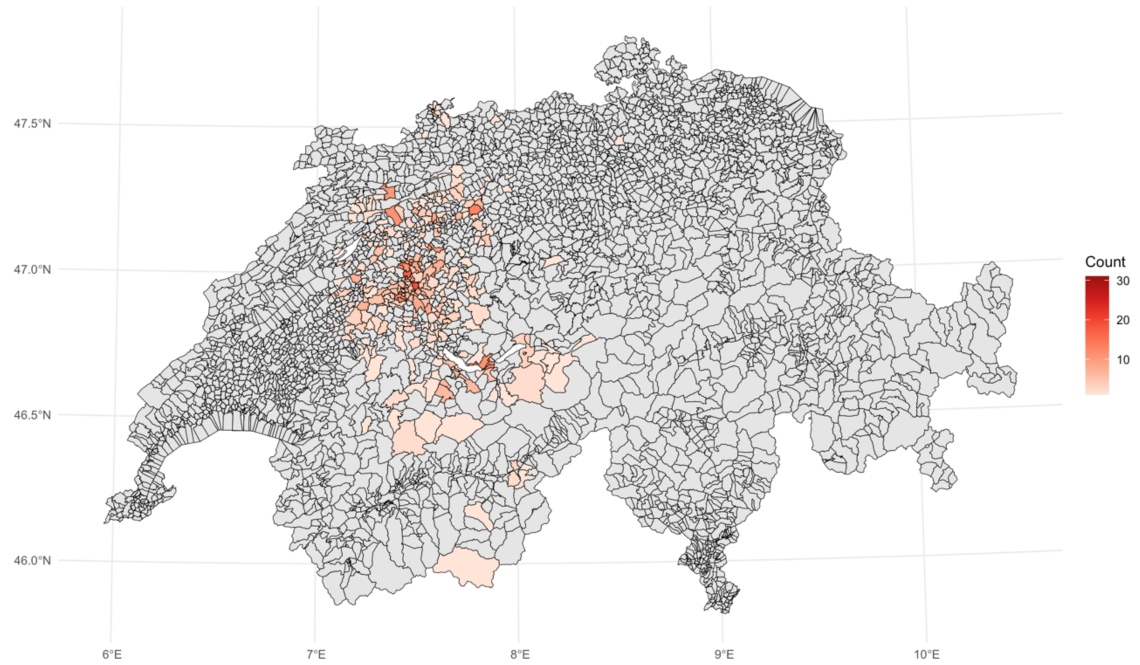


**Figure A.1** Geographic distribution of BOCAB participants across Switzerland according to postal code of residence at baseline visit. Shades of red indicate the number of participants per postal code area.

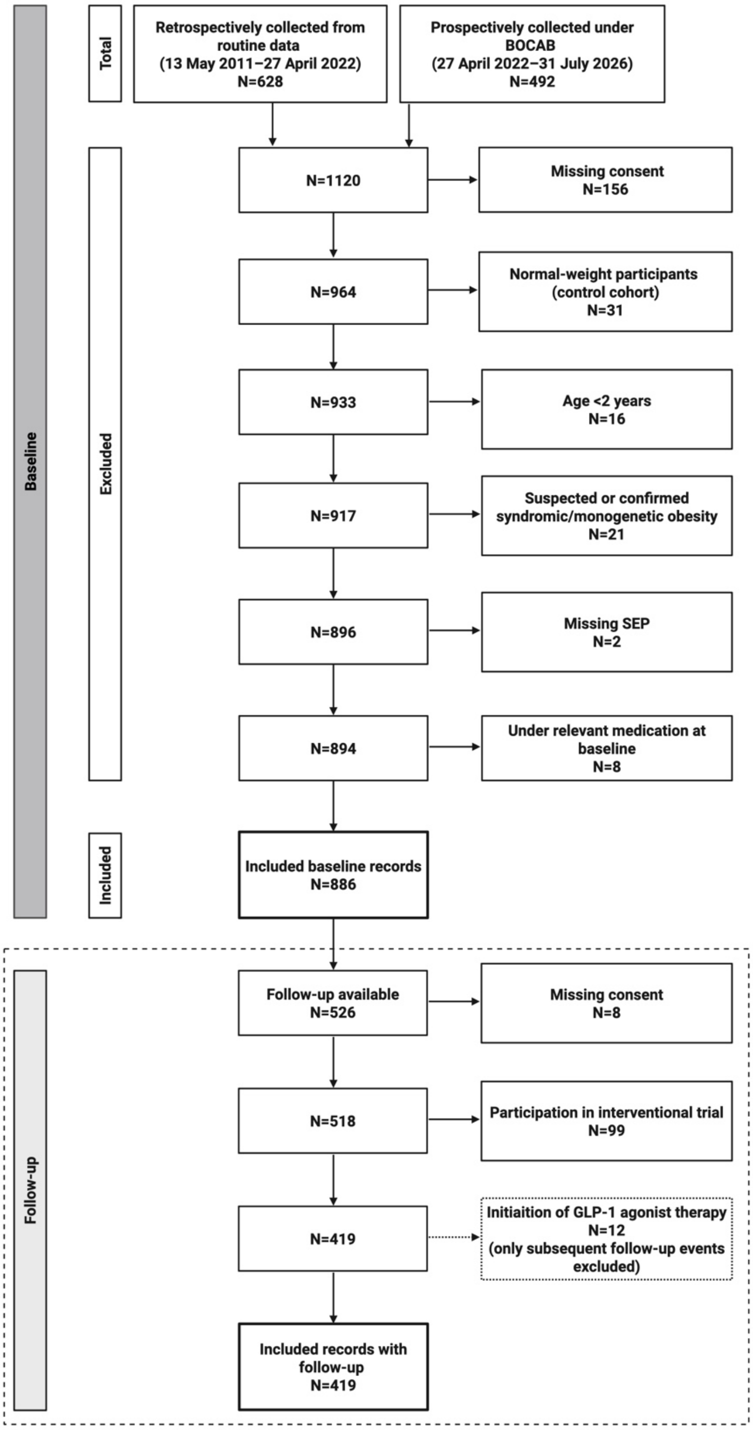


**Figure A.2** Flow diagram of patient inclusion and exclusion for the present study. At baseline, relevant medication refers to all outcome-modifying drugs (e.g., lipid-lowering drugs, antihypertensive drugs, metformin, GLP-1 receptor agonists). For follow-up events, only GLP-1 receptor agonist therapy was considered relevant for exclusion, as only weight outcomes were analyzed longitudinally in this study. SEP, socioeconomic position; GLP-1, glucagon-like peptide-1. Created in BioRender. Koch, A. (2026) https://BioRender.com/ty3bh77

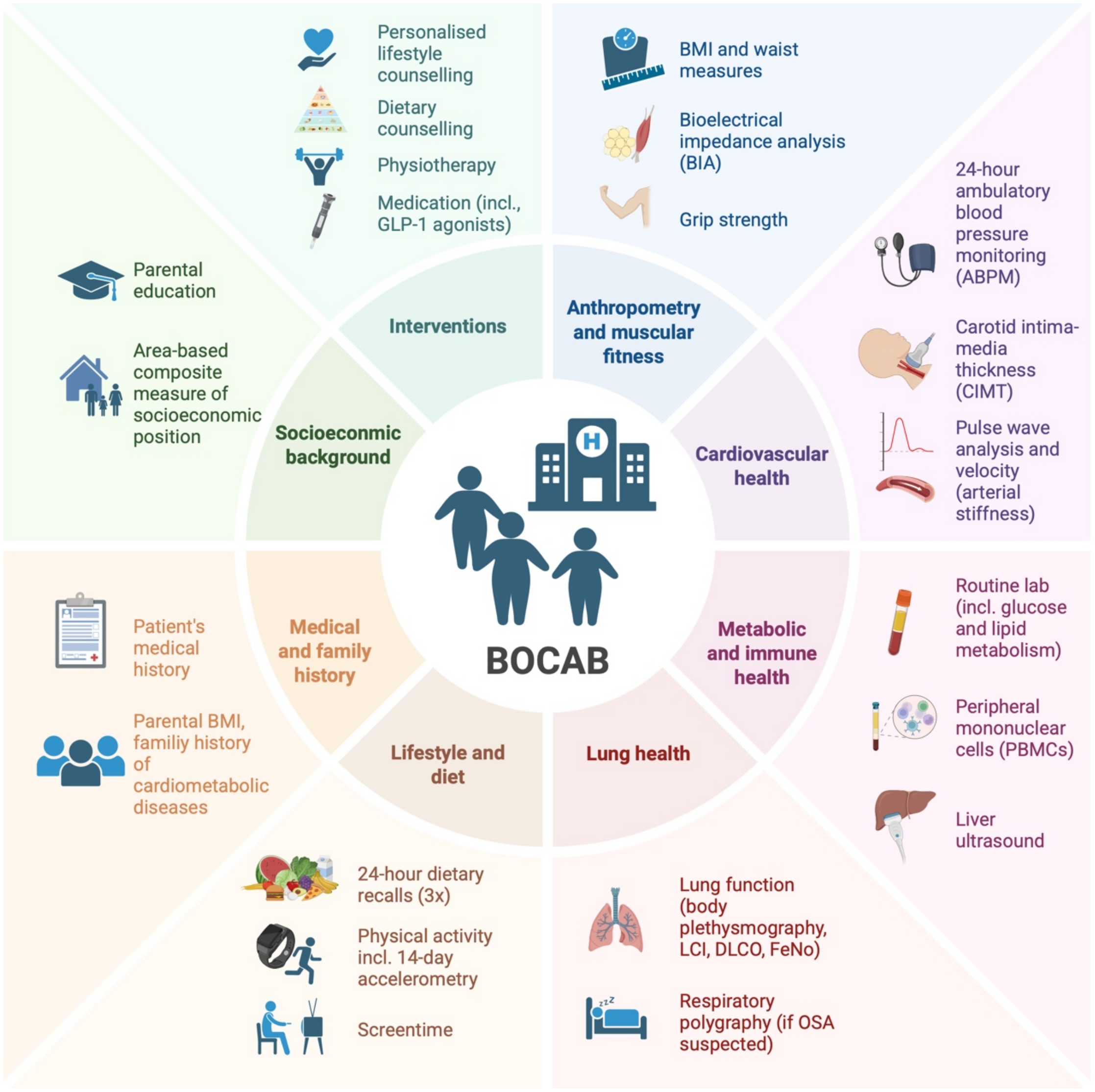


**Figure A.3** Summary of assessments and procedures implemented in the Bern Obesity in Childhood and Adolescence Biorepository (BOCAB) for comprehensive phenotyping. Routine assessments expanded after the weight management service launched in 2011; thus, data availability varies across participants and assessment types. For example, 24-hour ambulatory blood pressure monitoring was introduced in 2012, liver ultrasound in 2016 (for participants aged >10 years), bioelectric impedance analysis in 2018, grip strength measurement in 2022, biobank and pulse wave analysis/arterial stiffness assessment in 2023, lung function testing in 2024, dietary recalls in 2022, accelerometry in 2023, and CIMT in 2026. BMI, body mass index; GLP-1, glucagon-like peptide-1; LCI, lung clearance index; DLCO, diffusing capacity of the lung for carbon monoxide; FeNO, fractional exhaled nitric oxide; OSA, obstructive sleep apnea. Created in BioRender. Koch, A. (2026) https://BioRender.com/4ykzlnl

**Table A.1** Overview and schedule of routine and study-related assessments and procedures at the pediatric weight management service of the Bern University Children's Hospital. Adapted from BOCAB protocol version 5.0.

| Procedures/ Assessments | Control group[a] | Patients with obesity | | | |
|---|---|---|---|---|---|
| | Baseline | Baseline | 6 months | 12 months | Annually follow-up[d] |
| Eligibility check | X | X | | | |
| Written informed consent | X | X | | | |
| Clinical examination (incl. urine collection) | X | X | X | X | X |
| Bioelectric impedance analysis (BIA) | X | X | X | X | X |
| Grip strength assessment | X | X | X | X | X |
| 24-hour ambulatory blood pressure | X | X | X[b] | X[b] | X[b] |
| Puls wave analysis / arterial stiffness | X | X[b] | X[b] | X[b] | X[b] |
| Carotid intima-media thickness | X | X[b] | X[b] | X[b] | X[b] |
| Fasting venous blood sample[c] | X | X[b] | X[b] | X[b] | X[b] |
| Liver ultrasound | X | X | X[b] | X[b] | X[b] |
| Lung function testing | X | X[b] | | | |
| 24-hour dietary recalls (3x) | X | X | X[b] | X[b] | X[b] |
| 14-day wrist-based triaxial accelerometry | X | X | X[b] | X[b] | X[b] |
| Medical history | X | X | | | |
| Family history | X | X | | | |
| Parent questionnaire | X | X | | | |
| Child questionnaire | X | X | | | |

[a] For the control group, all procedures are study-related.

[b] Study-related procedures and assessments for patients with obesity attending the weight management service. Repeated assessments beyond baseline may depend on patient consent, clinical indication (e.g., previously abnormal values or substantial weight changes), available funding, and the predominant research questions at the time.

[c] The venous blood sample is routinely applied at baseline, but for the purpose of the study, the collected blood volume is higher for biobank storage.

[d] Annual follow-up assessments are planned up to age 17 or last visit.

### Extended methods: Cardiometabolic risk factor assessment

Office systolic and diastolic blood pressure (SBP/DBP) were measured in supine position after a 5-minute rest using a manual auscultatory device with an appropriately sized cuff [1]. SDS values were calculated based on reference data from National Heart, Lung, and Blood Institute (NHLBI) [2]. BP was categorized according to the 2023 European Society of Hypertension guidelines as follows: optimal, normal BP, high-normal BP, grade 1 hypertension, grade 2 hypertension and grade 3 hypertension, whereas optimal BP and grade 3 hypertension were only applicable for adolescents >16 years [1]. A binary variable was created classifying participants without hypertension (optimal, normal and high-normal BP) vs participants with hypertension (grade 1–3 hypertension).

24-h ambulatory blood pressure monitoring (ABPM) was performed on the right upper arm using validated oscillometric devices (Spacelabs OnTrak 90277 or 90217A, Spacelabs Healthcare Inc., Surrey, United Kingdom). BP measurements were obtained every 15 minutes during daytime and every 30 minutes during nighttime, with failed measurements repeated up to two times. Recordings were reviewed by a pediatric nephrologist using Sentinal software (version v11.5.7.17317, Spacelabs Healthcare Inc., Surrey, United Kingdom). Minimum quality criteria for inclusion were ≥20 valid daytime measurements and ≥7 valid nighttime measurements [3], and ≥70% valid measurements overall [4]. SDS values were calculated based on standard reference values published by the German Working Group on Pediatric Hypertension [5]. ABPM results were classified according to the 2016 European Society of Hypertension (ESH) pediatric guidelines [6], applying updated cut-off values from 2018 European Society of Cardiology (ESC)/ESH guideline [7]. Nocturnal dipping was calculated as the percentage decline mean SBP from daytime to nighttime, with non-dipping defined as nocturnal decline of less than 10% [6].

Venous blood samples were obtained after an overnight fast of at least 8-hours (see Online Resource 1, Table A.2 for detailed analytical methods). Continuous biomarkers were analyzed on their original scale, while predefined clinical or reference-based thresholds were used to derive binary cardiometabolic risk outcomes. Lipid profiles—including low-density lipoprotein cholesterol (LDL-C), high-density lipoprotein cholesterol (HDL-C), non-HDL-cholesterol (non-HDL-C) and triglycerides (TG)—were classified according to the 2018 American Heart Association guidelines [8]. Each lipid parameter was categorized as acceptable, borderline, or abnormal. Dyslipidemia was defined as the presence of at least one abnormal lipid value; participants were classified as having no dyslipidemia only when the complete lipid profile was available and all values were within reference. Glucose metabolism was assessed using fasting plasma glucose (FPG), HbA1c, and, where available, 2-hour plasma glucose (2hPG) during a 1.75g/kg bodyweight (max. 75-gram) oral glucose tolerance test (OGTT). Prediabetes and suspected T2DM were classified according to the 2024 ISPAD biochemical criteria as follows [9]: FPG 5.6–6.9 mmol/L (prediabetes) or ≥7.0 mmol/L (T2DM); HbA1c 5.7–6.4% (prediabetes) or: ≥6.5% (T2DM); or 2hPG 7.8–11.0 mmol/L (prediabetes) or ≥11.1 mmol/L (T2DM). As clinical symptoms of hyperglycaemia were not consistently available, prediabetes and suspected T2DM were combined into a binary outcome (no prediabetes/T2DM vs prediabetes/T2DM).

Homeostasis model assessment for insulin resistance (HOMA-IR) was calculated as fasting plasma glucose (FPG) [mmol/L] x fasting insulin [mU/L]) / 22.5. Insulin resistance and elevated fasting insulin were defined using pubertal-stage-specific, or age-specific where unavailable, 95th percentile thresholds from the LIFE Child cohort [10].

Alanine aminotransferase (ALT) was classified as elevated using sex-specific thresholds of >25.8 U/L for boys and >22.1 U/L for girls, based on NHANES-derived pediatric reference thresholds [11, 12].

**Table A.2** Extended methods for laboratory parameters used in the main text. All assays were performed according to the manufacturers' recommendations.

| **Lab parameter** | **Assay kit** | **Analytic method** |
|---|---|---|
| HDL-cholesterol | HDLC4, Roche Diagnostics, Mannheim, Germany; REF 07528582190 | Homogeneous enzymatic colorimetric assay; analyzed on cobas c platform |
| LDL-cholesterol | LDLC3, Roche Diagnostics, Mannheim, Germany; REF 07005717190 | Homogeneous enzymatic colorimetric assay; analyzed on COBAS INTEGRA platform |
| Triglycerides | TRIGL, Roche Diagnostics, Mannheim, Germany; REF 20767107322 | Enzymatic colorimetric assay; analyzed on cobas c or COBAS INTEGRA platforms |
| Glucose | GLUC3, Roche Diagnostics, Mannheim, Germany; REF 05168791190 | Enzymatic reference method with hexokinase; analyzed on cobas c platform |
| Insulin | Elecsys Insulin, Roche Diagnostics, Mannheim, Germany; REF 07027559190 | Electrochemiluminescence immunoassay (ECLIA) employing two monoclonal antibodies specific for human insulin; analyzed on cobas e platform |
| HbA1c | A1C-3, Roche Diagnostics, Mannheim, Germany; REF 05168791190 | Turbidimetric inhibition immunoassay (TINIA) for hemolyzed whole blood; analyzed on cobas c or COBAS INTEGRA platforms |
| | DCA HbA1c Reagin Kit, Siemens Healthcare Diagnostics Inc., Tarrytown, USA; REF 5035C | Latex immunoagglutination inhibition; analyzed on DCA Vantage platform |
| ALT | ALTPM, Roche Diagnostics, Mannheim, Germany; REF 05531462190 | Enzymatic UV kinetic assay with pyridoxal phosphate (P-5'-P) activation, analyzed on the cobas c platform |

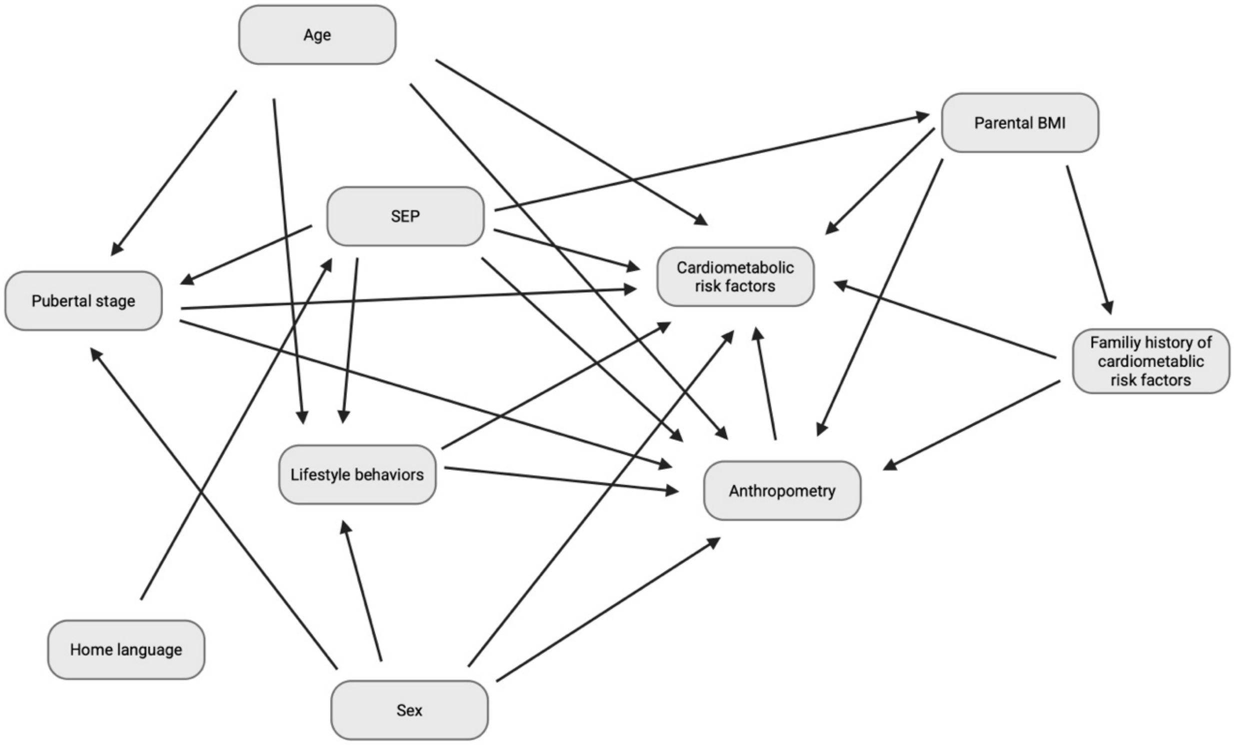


**Figure A.4** Directed acyclic graph (DAG) illustrating the hypothesized relationships between socioeconomic position (SEP), potential mediating pathways, confounders and anthropometric and cardiometabolic outcomes. Arrows represent hypothesized causal relationships and the assumed causal structure underlying the analyses. Unless otherwise specified, all variables refer to the child. Created in BioRender. Koch, A. (2026) https://BioRender.com/wh60ljp

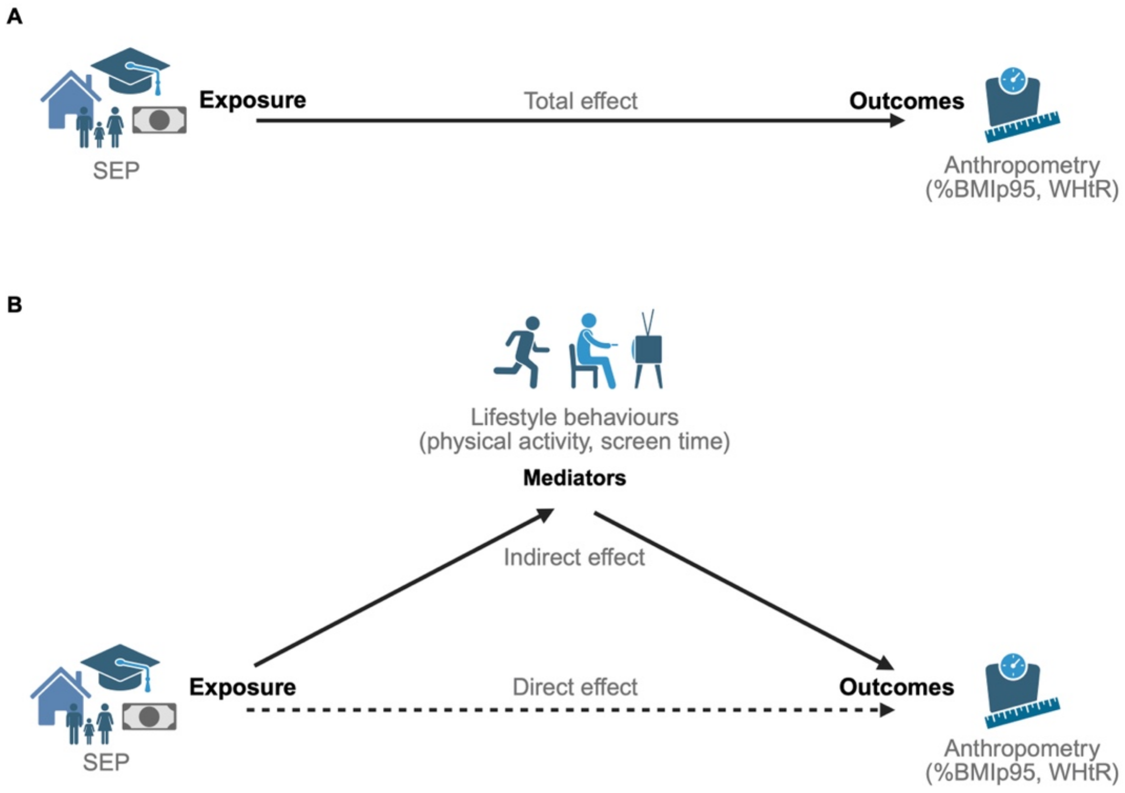


**Figure A.5** Mediation framework to explore mediators of the associations of socioeconomic position (SEP) with %BMIp95 and WHtR. Mediation analyses were conducted to investigate whether physical activity and screen time could account for part of the observed associations by decomposing the total effect (A) into an indirect effect (transmitted through mediator) and direct effect (residual effect without mediating pathway) (B). SEP, socioeconomic position; %BMIp95, percentage of the 95th BMI percentile for age and sex; WHtR, waist-to-height ratio. Created in BioRender. Koch, A. (2026) https://BioRender.com/4ykzlnl

### Supplementary results

**Table A.3** Baseline characteristics of participants with vs. without available follow-up.

| Characteristic | No follow-up (N=467) | With follow-up (N=419) | p |
|---|---|---|---|
| **Demographics** | | | |
| Age, years | 11.4 (3.0) | 11.0 (3.2) | **0.040** |
| Sex, males | 234 (50.1%) | 225 (53.7%) | 0.32 |
| Pubertal stage (Tanner) | [n=408] | [n=368] | 0.24 |
| Prepubertal | 177 (43.4%) | 181 (49.2%) | |
| Peripubertal | 123 (30.1%) | 95 (25.8%) | |
| Postpubertal | 108 (26.5%) | 92 (25.0%) | |
| SEP | 61.13 (8.89) | 60.97 (9.03) | 0.79 |
| Home language | [n=459] | [n=393] | 0.91 |
| Swiss (German/French/Italian) | 332 (72.3%) | 282 (71.8%) | |
| Other | 127 (27.7%) | 111 (28.2%) | |
| **Anthropometry & body composition** | | | |
| Height, cm | 152.4 (16.9) | 150.6 (18.9) | 0.13 |
| Weight, kg | 70.3 (24.3) | 70.2 (27.0) | 0.95 |
| BMI, kg/m2 | 29.2 (5.2) | 29.6 (5.7) | 0.30 |
| BMI SDS | 2.25 (0.59) | 2.39 (0.63) | **<0.001** |
| %BMIp95 | 122.0 (17.0) | 125.6 (17.7) | **0.0022** |
| WC, cm | 89.0 (12.8) [n=416] | 89.2 (14.2) [n=382] | 0.89 |
| WC SDS | 1.74 (0.42) [n=406] | 1.82 (0.40) [n=359] | **0.0037** |
| WHtR | 0.584 (0.055) [n=416] | 0.593 (0.059) [n=382] | **0.018** |
| %BF | 41.9 (6.5) [n=346] | 42.1 (6.6) [n=237] | 0.72 |
| %MM | 31.4 (3.6) [n=348] | 31.3 (3.6) [n=237] | 0.88 |
| **Blood pressure** | | | |
| Office measurements | | | |
| Office SBP, mmHg | 110 (11) [n=420] | 108 (12) [n=363] | 0.100 |
| Office DBP, mmHg | 70 (9) [n=418] | 68 (9) [n=363] | **<0.001** |
| 24-h ABPM | | | |
| 24-h SBP, mmHg | 109 (8) [n=197] | 110 (9) [n=210] | **0.034** |
| 24-h DBP, mmHg | 64 (5) [n=197] | 65 (5) [n=210] | **0.047** |
| **Lipids** | | | |

| Characteristic | No follow-up (N=467) | With follow-up (N=419) | p |
|---|---|---|---|
| HDL-C, mmol/L | 1.20 (0.26) [n=423] | 1.20 (0.27) [n=389] | 0.84 |
| LDL-C, mmol/L | 2.60 (0.73) [n=417] | 2.65 (0.71) [n=381] | 0.30 |
| Non-HDL-C, mmol/L | 2.90 (0.80) [n=422] | 2.91 (0.76) [n=389] | 0.96 |
| TG, mmol/L | 1.16 (0.69) [n=422] | 1.16 (0.60) [n=389] | 0.97 |
| **Metabolism & liver** | | | |
| FPG, mmol/L | 5.10 (0.44) [n=429] | 5.11 (0.51) [n=397] | 0.63 |
| Fasting insulin, mU/L | 23.5 (18.0) [n=341] | 20.6 (12.8) [n=206] | **0.033** |
| HOMA-IR | 5.44 (4.62) [n=340] | 4.69 (3.51) [n=205] | **0.032** |
| 2-h PG, mmol/L | 6.40 (1.37) [n=270] | 6.46 (1.40) [n=222] | 0.64 |
| HbA1c, % | 5.4 (0.3) [n=260] | 5.5 (0.3) [n=88] | **0.044** |
| ALT, U/L | 25 (13) [n=424] | 31 (26) [n=395] | **<0.001** |
| **Lifestyle & family** | | | |
| Physical activity, h/week | 3.5 (2.5) [n=425] | 3.5 (2.6) [n=388] | 0.70 |
| Screentime, h/week | 18.1 (12.5) [n=415] | 17.3 (12.4) [n=382] | 0.34 |
| Parental BMI | | | |
| Maternal BMI, kg/m2 | 28.9 (6.6) [n=351] | 28.8 (6.3) [n=320] | 0.89 |
| Paternal BMI, kg/m2 | 28.8 (4.8) [n=331] | 29.6 (5.2) [n=277] | 0.078 |
| Family history of | | | |
| T2DM | 200 (48.5%) [n=412] | 185 (50.4%) [n=367] | 0.65 |
| Hypertension | 250 (59.8%) [n=418] | 204 (55.4%) [n=368] | 0.24 |
| Dyslipidemia | 148 (36.5%) [n=406] | 107 (30.0%) [n=357] | 0.069 |

Note. All characteristics measured at baseline for both columns. Continuous variables: mean (SD), with N in brackets where less than the column total, compared with Welch's t-test. Categorical variables: n (%) of non-missing, compared with chi-squared or Fisher's exact test.

SEP, socioeconomic position; BMI, body mass index; SDS, standard deviation score; %BMIp95, percentage of the 95th BMI percentile; WC, waist circumference; WHtR, waist-to-height ratio; %BF, percentage body fat; %MM, percentage muscle mass; SBP, systolic blood pressure; DBP, diastolic blood pressure; ABPM, ambulatory blood pressure monitoring; HDL-C, high-density lipoprotein cholesterol; LDL-C, low-density lipoprotein cholesterol; non-HDL-C, non-high-density lipoprotein cholesterol; TG, triglycerides; FPG, fasting plasma glucose; HOMA-IR, homeostatic model assessment for insulin resistance; 2hPG, 2-hour plasma glucose; HbA1c, glycated hemoglobin; ALT, alanine aminotransferase; T2DM, type 2 diabetes mellitus

**Table A.4** Adjusted linear regression models of the associations of socioeconomic position (SEP) with anthropometry and cardiometabolic risk factors. Effect estimates per 10-unit higher SEP.

| | Model A (age, sex, puberty)[b] | | | Model B (age, sex, language) | | | Model C (age, sex, language, parental BMI, ±family history)[c] | | |
|---|---|---|---|---|---|---|---|---|---|
| **Outcome** | **β (95% CI)** | **p** | **n[a]** | **β (95% CI)** | **p** | **n[a]** | **β (95% CI)** | **p** | **n[a]** |
| **Anthropometry & body composition** | | | | | | | | | |
| BMI, kg/m² | -0.64 (-0.97, -0.32) | **<0.001** | 776 | -0.56 (-0.87, -0.24) | **<0.001** | 852 | -0.50 (-0.86, -0.14) | **0.0066** | 570 |
| BMI SDS | -0.09 (-0.14, -0.04) | **<0.001** | 776 | -0.08 (-0.12, -0.04) | **<0.001** | 852 | -0.07 (-0.13, -0.02) | **0.0081** | 570 |
| %BMIp95 | -2.77 (-4.12, -1.43) | **<0.001** | 776 | -2.39 (-3.68, -1.10) | **<0.001** | 852 | -2.23 (-3.73, -0.74) | **0.0034** | 570 |
| WC, cm | -1.38 (-2.14, -0.62) | **<0.001** | 715 | -1.16 (-1.91, -0.42) | **0.0022** | 773 | -1.04 (-1.88, -0.20) | **0.016** | 528 |
| WC SDS | -0.06 (-0.09, -0.03) | **<0.001** | 682 | -0.05 (-0.08, -0.02) | **0.0015** | 741 | -0.05 (-0.09, -0.01) | **0.012** | 502 |
| WHtR | -0.007 (-0.011, -0.002) | **0.0051** | 715 | -0.006 (-0.010, -0.002) | **0.0090** | 773 | -0.005 (-0.010, 0.000) | 0.059 | 528 |
| %BF | -1.09 (-1.71, -0.47) | **<0.001** | 521 | -0.71 (-1.29, -0.13) | **0.017** | 557 | -0.63 (-1.29, 0.03) | 0.061 | 413 |
| %MM | 0.61 (0.27, 0.96) | **<0.001** | 523 | 0.40 (0.07, 0.73) | **0.017** | 559 | 0.35 (-0.03, 0.73) | 0.070 | 414 |
| **Blood pressure** | | | | | | | | | |
| Office measurements | | | | | | | | | |
| Office SBP SDS | -0.05 (-0.13, 0.03) | 0.18 | 706 | -0.05 (-0.13, 0.02) | 0.18 | 758 | -0.04 (-0.14, 0.05) | 0.37 | 507 |
| Office DBP SDS | -0.01 (-0.07, 0.06) | 0.84 | 704 | -0.00 (-0.06, 0.06) | 0.98 | 756 | 0.01 (-0.07, 0.08) | 0.85 | 505 |
| 24-h ABPM | | | | | | | | | |
| 24h SBP SDS | -0.09 (-0.22, 0.04) | 0.16 | 362 | -0.09 (-0.21, 0.02) | 0.12 | 394 | -0.08 (-0.21, 0.06) | 0.28 | 294 |
| 24h DBP SDS | -0.08 (-0.18, 0.02) | 0.11 | 362 | -0.11 (-0.20, -0.01) | 0.025 | 394 | -0.08 (-0.19, 0.03) | 0.134 | 294 |
| **Lipids** | | | | | | | | | |

| | Model A (age, sex, puberty)[b] | | | Model B (age, sex, language) | | | Model C (age, sex, language, parental BMI, ±family history)[c] | | |
|---|---|---|---|---|---|---|---|---|---|
| **Outcome** | **β (95% CI)** | **p** | **n[a]** | **β (95% CI)** | **p** | **n[a]** | **β (95% CI)** | **p** | **n[a]** |
| HDL-C, mmol/L | 0.01 (-0.01, 0.03) | 0.29 | 718 | 0.01 (-0.01, 0.03) | 0.38 | 782 | -0.00 (-0.03, 0.03) | 0.985 | 494 |
| LDL-C, mmol/L | -0.04 (-0.09, 0.02) | 0.24 | 710 | -0.02 (-0.07, 0.04) | 0.60 | 768 | -0.02 (-0.09, 0.05) | 0.531 | 491 |
| Non-HDL-C, mmol/L | -0.04 (-0.11, 0.02) | 0.16 | 717 | -0.02 (-0.09, 0.04) | 0.43 | 781 | -0.03 (-0.10, 0.05) | 0.462 | 493 |
| TG, mmol/L | -0.03 (-0.08, 0.02) | 0.24 | 717 | -0.03 (-0.08, 0.02) | 0.21 | 781 | -0.02 (-0.08, 0.04) | 0.511 | 494 |
| **Metabolism & liver** | | | | | | | | | |
| FPG, mmol/L | 0.00 (-0.04, 0.04) | 0.96 | 730 | -0.01 (-0.04, 0.03) | 0.77 | 796 | 0.00 (-0.04, 0.05) | 0.97 | 515 |
| Fasting insulin, mU/L | -1.65 (-3.12, -0.17) | **0.029** | 484 | -1.38 (-2.87, 0.10) | 0.068 | 534 | -1.34 (-3.50, 0.82) | 0.22 | 324 |
| HOMA-IR | -0.39 (-0.78, -0.01) | **0.045** | 482 | -0.32 (-0.72, 0.07) | 0.10 | 532 | -0.33 (-0.92, 0.26) | 0.27 | 322 |
| 2-h PG, mmol/L | -0.05 (-0.20, 0.10) | 0.50 | 419 | -0.02 (-0.16, 0.12) | 0.78 | 475 | -0.03 (-0.22, 0.17) | 0.79 | 300 |
| HbA1c, % | -0.03 (-0.07, 0.01) | 0.16 | 315 | -0.03 (-0.07, 0.01) | 0.18 | 333 | -0.01 (-0.06, 0.04) | 0.66 | 242 |
| ALT, U/L | -0.41 (-2.14, 1.31) | 0.64 | 725 | -0.43 (-2.04, 1.19) | 0.60 | 790 | 0.39 (-1.20, 1.97) | 0.63 | 532 |
| **Lifestyle** | | | | | | | | | |
| Physical activity, h/week | 0.28 (0.07, 0.48) | **0.0087** | 717 | 0.22 (0.02, 0.41) | **0.029** | 785 | 0.16 (-0.08, 0.41) | 0.18 | 541 |
| Screentime, h/week | -1.05 (-1.95, -0.15) | **0.023** | 706 | -0.93 (-1.79, -0.07) | **0.034** | 768 | -1.19 (-2.22, -0.17) | **0.023** | 530 |

[a]Sample size reflects complete cases for that model; may vary across models due to missing covariate data.

[b]Per the DAG, pubertal stage is a hypothesized mediator (SEP may influence pubertal timing) rather than a confounder of the SEP-outcome association; this model therefore estimates a direct effect not operating through pubertal stage, rather than a more completely confounder-adjusted total effect. The cross-sectional measurement of anthropometric outcomes and pubertal stage (same visit) means the temporal ordering between them could not be established, and some bidirectionality is biologically plausible.

[c]Additionally adjusted for maternal and paternal BMI and, where clinically relevant, family history (T2DM for glycaemic outcomes; dyslipidemia for lipid outcomes; hypertension for blood pressure outcomes). Per the directed acyclic graph (Section 12), parental BMI is a hypothesized mediator rather than a confounder of the SEP-outcome association; this model therefore estimates a direct effect not operating through parental BMI, rather than a more completely confounder-adjusted total effect.

| | Model A (age, sex, puberty)[b] | | | Model B (age, sex, language) | | | Model C (age, sex, language, parental BMI, ±family history)[c] | | |
|---|---|---|---|---|---|---|---|---|---|
| **Outcome** | **β (95% CI)** | **p** | **n[a]** | **β (95% CI)** | **p** | **n[a]** | **β (95% CI)** | **p** | **n[a]** |

SEP, socioeconomic position; CI, confidence interval; BMI, body mass index; SDS, standard deviation score; %BMIp95, percentage of the 95th BMI percentile; WC, waist circumference; WHtR, waist-to-height ratio; %BF, percentage body fat; %MM, percentage muscle mass; SBP, systolic blood pressure; DBP, diastolic blood pressure; ABPM, ambulatory blood pressure monitoring; HDL-C, high-density lipoprotein cholesterol; LDL-C, low-density lipoprotein cholesterol; non-HDL-C, non-high-density lipoprotein cholesterol; TG, triglycerides; FPG, fasting plasma glucose; HOMA-IR, homeostatic model assessment for insulin resistance; 2hPG, 2-hour plasma glucose; HbA1c, glycated hemoglobin; ALT, alanine aminotransferase; T2DM, type 2 diabetes mellitus

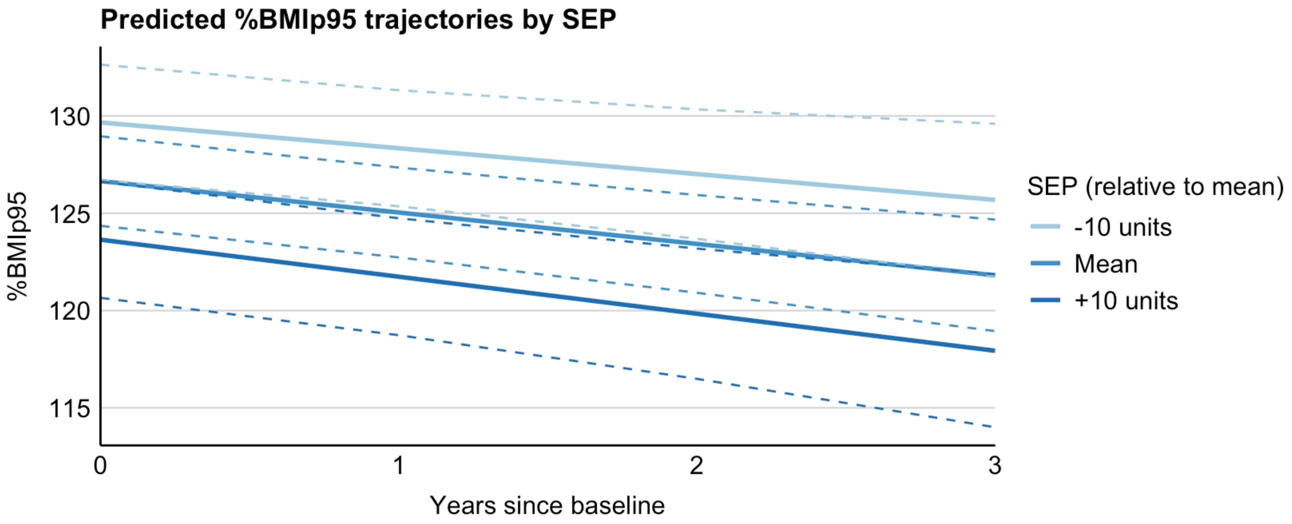


**Figure A.6** Predicted %BMIp95 trajectories by socioeconomic position (SEP). Dashed lines represent 95% confidence intervals. %BMIp95, percentage of the 95th BMI percentile for age and sex; SEP, socioeconomic position.
